# Using self-controlled case series to understand the relationship between conflict and cholera in Nigeria and the Democratic Republic of Congo

**DOI:** 10.1101/2021.10.19.21265191

**Authors:** Gina E C Charnley, Kévin Jean, Ilan Kelman, Katy A M Gaythorpe, Kris A Murray

## Abstract

Cholera outbreaks significantly contribute to disease mortality and morbidity in low- and middle-income countries. Cholera outbreaks have several social and environmental risk factors and extreme conditions can act as catalysts. A social extreme with known links to infectious disease outbreaks is conflict, causing disruption to services, loss of income and displacement. Here, we used the self-controlled case series method in a novel application and found that conflict increased the risk of cholera in Nigeria by 3.6 times and 19.7% of cholera outbreaks were attributable to conflict. In the Democratic Republic of Congo (DRC), conflict increased the risk of cholera by 2.6 times and 12.3% of cholera outbreaks were attributable to conflict. Our results highlight the importance of rapid and sufficient assistance during conflict-related cholera outbreaks, while also working towards conflict resolution and addressing pre-existing vulnerabilities such as poverty and access to healthcare.

**Article Summary Line:** Conflict significantly increased the risk of cholera outbreaks in Nigeria and the Democratic Republic of Congo and pre-existing vulnerabilities and conflict resolution should be a top priority to protect health.

## Text

Diarrhoeal diseases are the eighth leading cause of death worldwide, with cholera contributing significantly, especially in low- and middle-income countries (*1*). Over 94% of World Health Organization (WHO) reported cholera cases are in Africa and more research is needed to understand cholera dynamics on the continent (*2*). Previous research has found several environmental and socioeconomic links with cholera, including temperature, precipitation, poverty and water, sanitation, and hygiene (WASH) (*3,4*). Furthermore, extremes of these environmental and social conditions can act as catalysts for outbreaks, such as droughts, floods, and conflicts (*4,5,6*).

Here, we will focus on the impacts of conflict on cholera outbreaks and compare the results across two countries in Africa, Nigeria and the Democratic Republic of Congo (DRC) in the past 23 years. Several mechanisms have been suggested through which conflict can lead to infectious disease outbreaks (*7,8,9*). During conflicts, services can be disrupted including access to WASH, disruption of disease control programmes and collapse of health systems (e.g., vaccination coverage). Those displaced by conflict may also find it difficult to access healthcare (*10,11,12*).

Populations may not seek medical treatment as they perceive healthcare facilities as unsafe. For example, during the 2018 Ebola outbreak in the DRC healthcare facilities were attacked, dampening efforts to control the virus (*12*). Conflict can worsen pre-existing vulnerabilities including poverty, as conflicts can cause loss of income, disruption to education, damage to livelihoods and displacement (*13*).

Nigeria and the DRC share social and environmental similarities, as well as experiencing cholera outbreaks. Both have active conflicts including the Boko Haram Insurgency in northeastern Nigeria (*14*) and political unrest in the eastern DRC (*15*). They have the second and third highest numbers of estimated cholera cases per year in Africa, respectively (*16*), with the Kivu provinces being the most active cholera foci in the world (*17*). In addition, Nigeria and the DRC have a tropical climate, poor access to WASH and a large proportion of the population living in poverty (<$1.25/day) at 87.7% for the DRC and 62% for Nigeria (*18*), which are known cholera risk factors.

Few studies have investigated the impacts of conflict on cholera outbreaks, especially quantitatively. Studies have commonly focused on cholera and conflict in Yemen (*8,19*), its effect on vaccination efforts (*20*) or the impact of conflict on other diseases such as Ebola (*12*) and COVID-19 (*21*). Africa is also a chronically understudied continent in relation to cholera, despite reporting a large proportion of global cases (*2*).

To bridge this research gap, we used the Self-Controlled Case Series (SCCS) method, both nationally and sub-nationally and completed a sensitivity analysis to provide insight into the effect of lag and cholera definition. The SCCS method is used in a novel application and we aim to understand and promote its use in other contexts (*22*). Previous uses of the method include testing the effectiveness of drug and vaccine intervention on an individual (*23,24*) and population level (*25*). Furthermore, we adapted the recently developed percentage attributable fraction (PAF) equations to the work presented here (*25*), to understand the proportion of cholera outbreaks attributable to conflict. Based on these results, we suggest mechanisms for which conflict is driving cholera and potential risk factors, building on previous research in this area. We hope this information can be used to strengthen disease prevention in conflict settings and reduce additional mortality and morbidity in conflicts.

## Methods

### Datasets

Cholera data were compiled from a range of publicly available sources (WHO disease outbreak news, ProMED, ReliefWeb, WHO regional office for Africa weekly outbreak and emergencies, UNICEF cholera platform, EM-DAT, the Nigerian Centre for Disease Control, and a literature search) in both English and French. The full compiled dataset is available in a GitHub repository (https://github.com/GinaCharnley/cholera_data_drc_nga) and additional information on data collation and validation in a complementary database paper (*26*). An outbreak was defined by the onset of a cholera case and the case definitions for the two countries are shown in S1 Information. Conflict data were provided by the United Nations Office for the Coordination of Humanitarian Affairs Humanitarian Data Exchange, which provides data from the Armed Conflct Location & Event Data Project (ACLED) (*27*). The data included sub-national conflict events, categorised by event type including battles, explosions, protests, riots, strategic developments, and violence against civilians.

The spatial granularity of the analysis was to administrative level 1 (states for Nigeria and provinces for the DRC) and all data points that were reported on a finer spatial scale were aggregated to the upper level. The study period was specified as January 1997 to May 2020, as these were the first and last reports in the conflict data. The temporal scale was set to weekly, with continuous weeks from epidemiological week 1 in 1997 to epidemiological week 20 in 2020 (1-1,220). Continuous weeks was chosen for compatibility with the model and to include periods of conflict that endured from one year into the next. Weeks was chosen, rather than days, to account for reporting lags, as previous work has reported issues in the granularity of data and timeliness of reporting, especially in humanitarian crises due to different sources of data and logistical difficulties (*28,29*). Additional information on the datasets used here are available in S2 Information.

### Model Structure and Fitting

The SCCS method investigates the association between an exposure and an outcome event. The aim is to estimate the effect, by comparing the relative incidence of the adverse events (outbreaks) within an exposure period of hypothesised excess risk (conflicts), compared to all other times (peace, according to the dataset used). The SCCS method is a case only method and has the advantage of not needing separate controls, by automatically controlling for fixed cofounders that remain constant over the observational period (*30,31*).

Both the event and exposure were set as binary outcomes, either being present (1) or not (0). The observation period was the full study period (1-1,220). The exposure period was the first week after conflict onset and was reported as multiple onsets for each event, not one long exposure period incorporating all events in the specific week (or 2, 4, 6, 8 and 10 weeks). The event was defined by the week the cholera outbreaks was reported. Each event and exposure that occurred in the same state/province were designated an identification number and a pre-exposure, exposure, and post exposure period (see S1 Table for data setup and additional information).

The data were fit to conditional logistic regression models, using the event (cholera outbreak onset) as the outcome variable (function clogit(), R package “survival”) (*32*). As is standard for conditional logistic regression, the interval between the exposure to un-exposure period was offset (coefficient value of 1) in the model and the identification numbers were stratified. The model coefficient values were used to calculate incidence rate ratio (IRR), which quantifies the magnitude in which conflict increased the rate of cholera outbreaks.

The datasets for each country were then split by state/province and the analysis repeated for each, to understand if the significance of conflict on cholera outbreaks varied by sub-national location and if conflict was more important in some states/provinces compared to others. All statistical analyses were carried out in R version 3.6.2 and the threshold for significance was p=<0.05.

### Sensitivity Analysis

A sensitivity analysis was used to test different methods of defining the exposure end point, which was set to one week in the main analysis, and 2, 4, 6, 8 and 10 weeks in the sensitivity analysis. The aim was to further understand how long after a conflict event the rate of cholera was heightened. Full information is available in S3 Information and S1 and S2 Figs.

An additional sensitivity analysis was completed to understand the effect of altering the cholera outbreak definition and to test for the presence of temporal autocorrelation. The analysis involved two scenarios. Scenario 1 removed all outbreaks within 2 weeks of each other (based on cholera biology, <10 days shedding the bacteria + <5 days incubation period) (*33,34*). Scenario 2 was an extreme scenario to fully test model robustness and removed all outbreaks within 6 months of each other.

### Percentage Attributable Fraction

The recently developed percentage attributable fraction (PAF) equations (*30*) were adapted to the model output and data (full equations in S4 Information). The PAF values estimate the percentage of outbreaks that could be attributed to conflict at a national level, using the full observation period of the datasets and the IRR values from the model results. Bootstrap resampling (1,000 samples) was used to obtain 95% confidence intervals. For each sample, a value of IRR was randomly sampled based on the parameters estimated in the SCCS analysis.

## Results

### Conflict and Cholera Occurrence

The distribution of conflict and cholera in Nigeria and the DRC in the datasets used here are shown temporally and spatially in Figure 1. The data show an increase in reported conflict and cholera, especially after 2010 (Figure 1a-d) and a large proportion of the cholera cases have been reported in conflict-stricken areas (Figure 1e-f).

**Figure 1.**
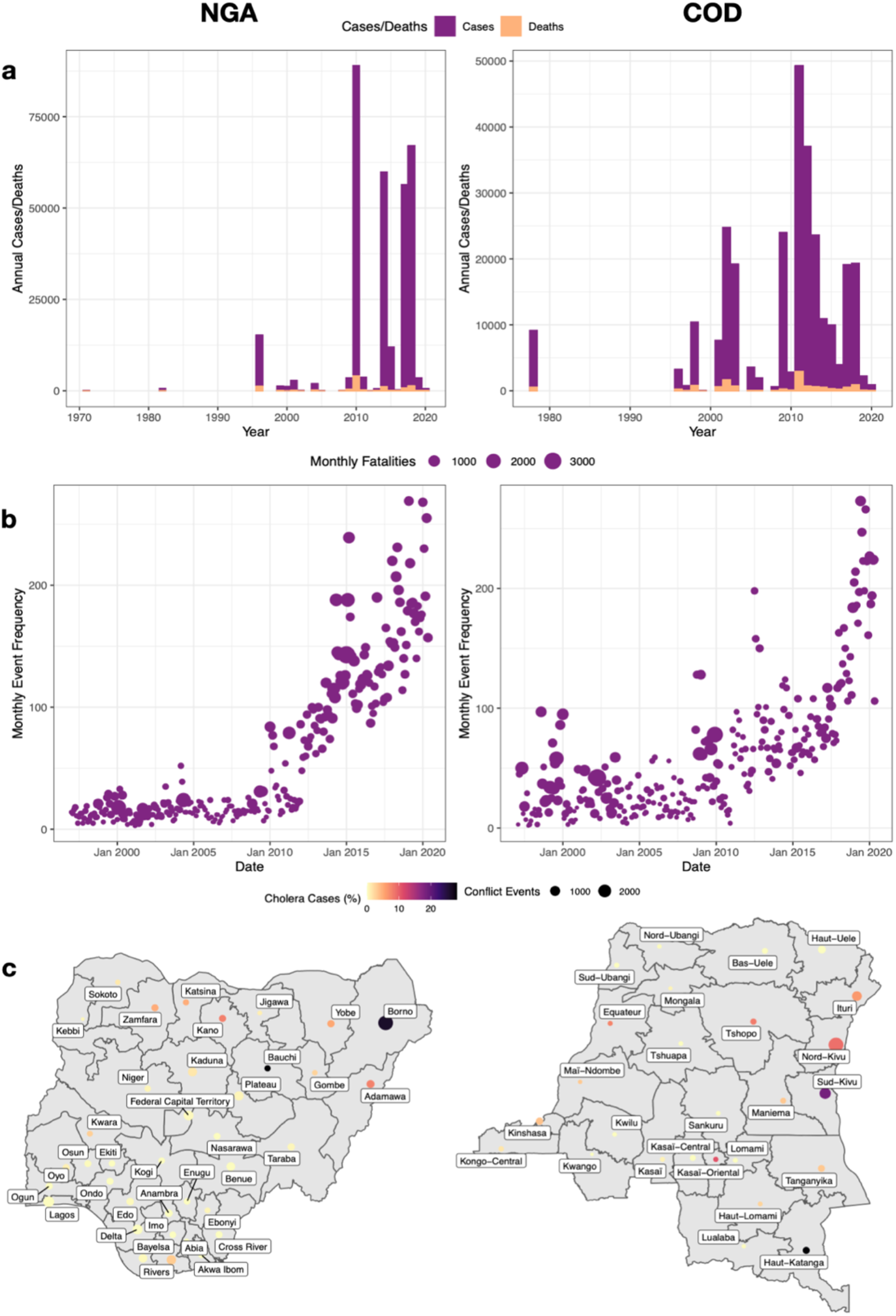
Changes in cholera and conflict for the full datasets. **a**, monthly cholera cases and deaths for Nigeria and **b**, the Democratic Republic of Congo. **c**, Monthly frequency of conflict events and fatalities for Nigeria and d, the Democratic Republic of Congo and **e** the number of conflict events and cholera cases as a percentage of the total number of national cases by administrative level 1 for Nigeria and **f**, the Democratic Republic of Congo.

The total number of conflicts and outbreaks for each state/province during the study period are shown below in Figure 2 and totaled 4,639 conflict and 396 cholera outbreaks for the DRC and 8,190 conflicts and 782 cholera outbreaks for Nigeria. The outbreak distribution applied satisfactorily to the Poisson probability distribution (S3 Figure).

**Figure 2.**
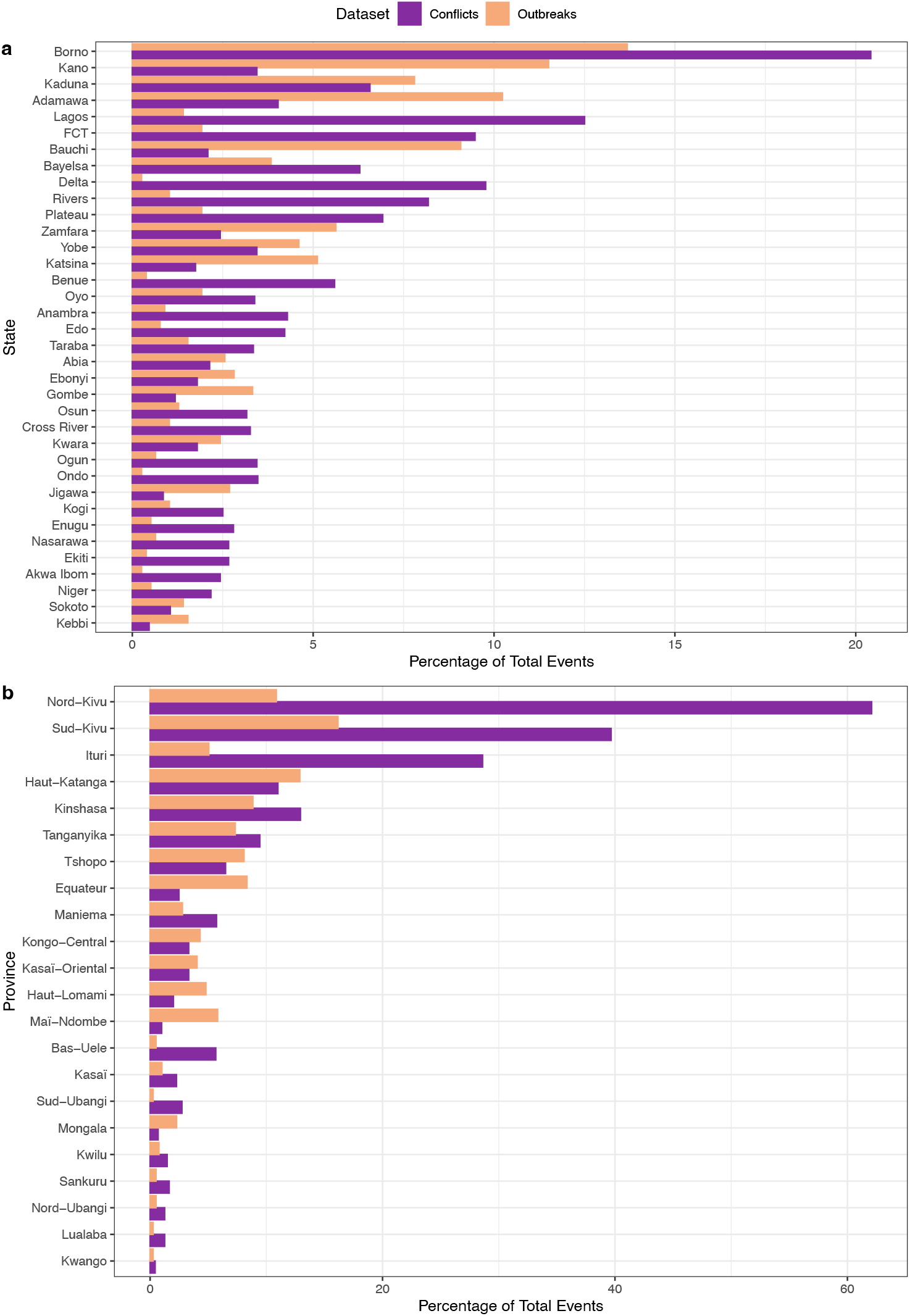
Percentage of events in each dataset including conflict and outbreaks, for **a**, Nigeria and **b**, the Democratic Republic of Congo by administrative level 1. FCT - Federal Capital Territory.

To be included in the analysis, the state/province had to report both outbreaks and conflicts during the study period, states/provinces which exclusively reported conflicts (and not any outbreaks) were excluded, as the SCCS methodology is a case-only approach. As such, 22 provinces were included for the DRC and 36 states for Nigeria (states and provinces excluded are shown in S5 Information). The temporal distribution of the exposure periods and outbreaks included in the SCCS model for each state/province are shown in Figure 3.

**Figure 3.**
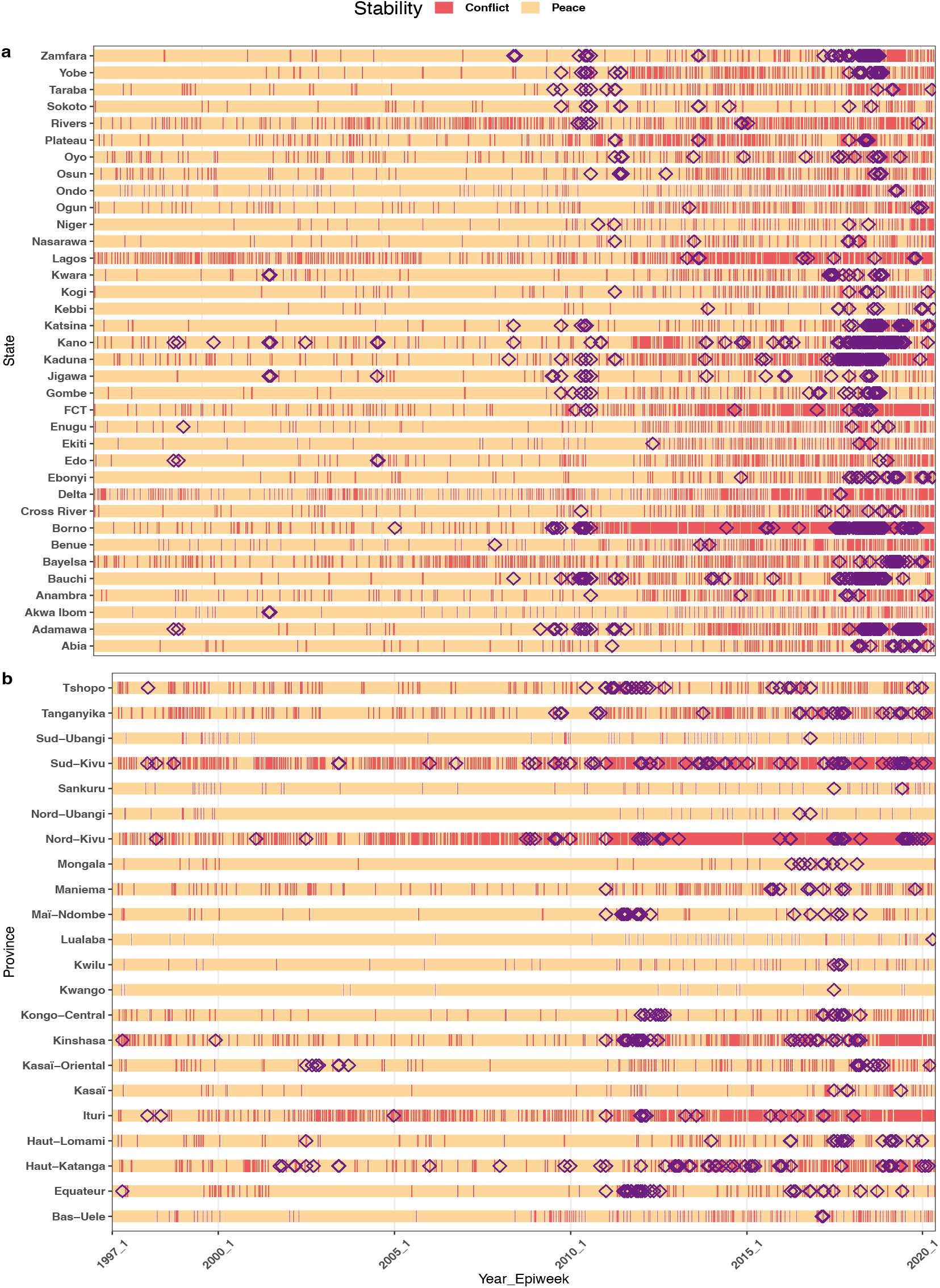
Swimmer plots showing the conflict exposure period in the SCCS model (1 week after the onset) and the outbreaks (black triangles) for each state/province for **a**, Nigeria and **b**, the Democratic Republic of Congo.

### Model Output

Conflict significantly increased the rate of cholera outbreaks (IRR) in the past 23 years in Nigeria and the DRC (p=<0.05). Nigeria had an effect of greater magnitude, increasing the risk of cholera outbreaks by up to 3.6 times (IRR = 3.6, 95%CI = 3.3-3.9). Whereas for the DRC, the risk was increased by 2.6 times (IRR = 2.6, 95%CI = 2.3-2.9).

Of the 36 Nigerian states included in the analysis, 24 showed statistically significant associations between conflict and cholera outbreaks. The strongest effect was found in Kebbi, Lagos, Osun, Borno and Nasarawa, with IRR values ranging from 6.8 to 6.2 (Figure 4a).

**Figure 4.**
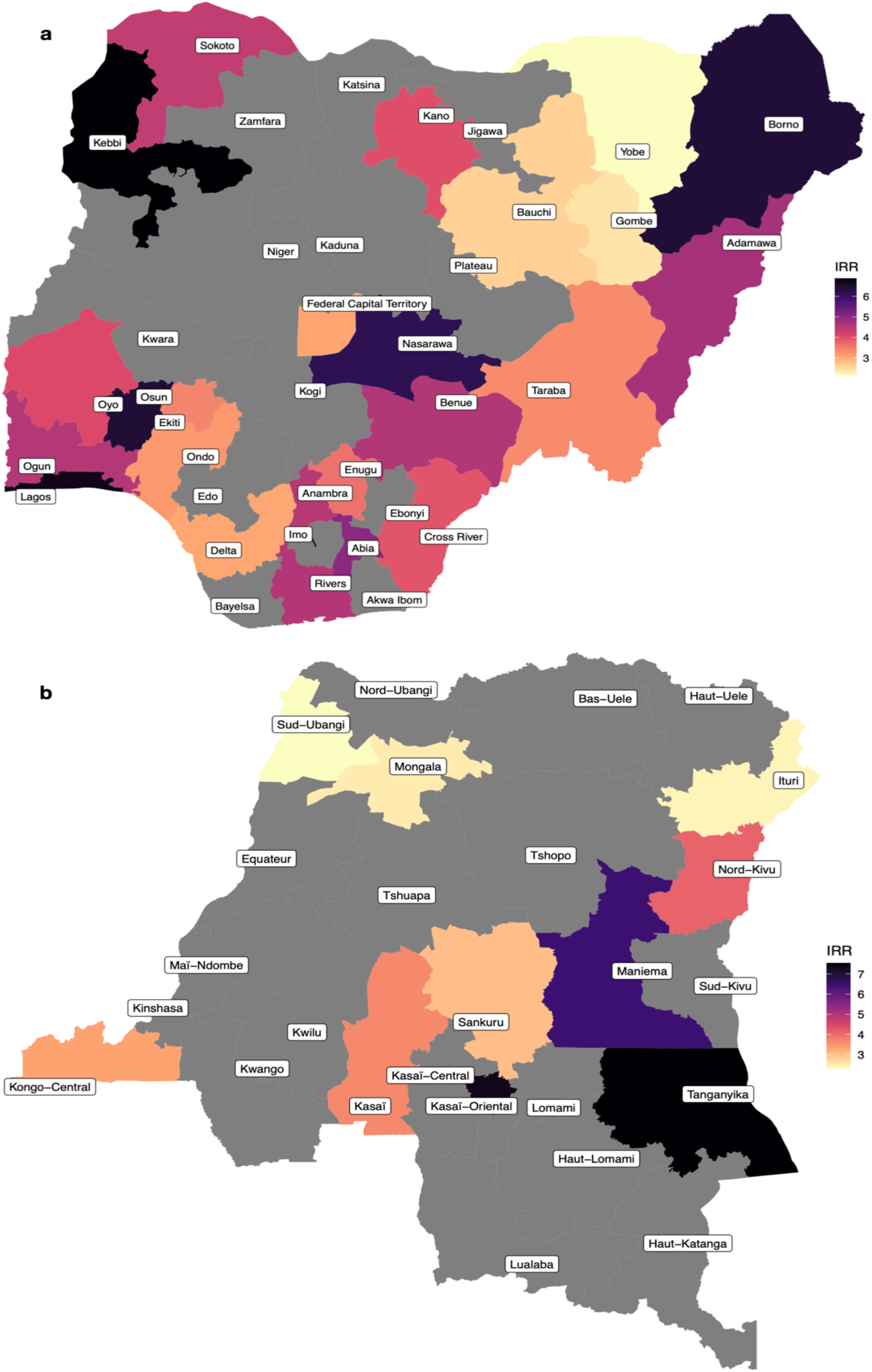
Incidence rate ratio (IRR) for the effect of exposure to conflict within one week of the event and cholera at a sub-national level. For **a**, Nigeria and **b**, the Democratic Republic of Congo. Only results that were significant at the threshold p=<0.05 are plotted here.

Eleven out of 22 DRC provinces included in the analysis showed a statistically significant relationship between conflict and cholera. Tanganyika, Kasaï-Oriental, Maniema, Nord-Kivu and Kasaï found the strongest values and some were the highest values found in the analysis. In Tanganyika, conflict increased cholera outbreak rate by 7.5 times and 3.7 times for Kasaï (Figure 4b).

### Sensitivity Analysis

The effect of conflict on cholera outbreaks at a national and sub-national level for both Nigeria and the DRC decreased with increasing exposure period. The decrease in IRR from week 1 to week 10 was from 3.6 to 2.08 for Nigeria and 2.6 to 1.5 for the DRC. By week 6 the change was minimal and plateaued or increased (full results in S4 and S5 Figs).

Changing the outbreak onset definition yielded similar results to the original analysis. Removing events within both 2 weeks and 6 months of each other found IRR values within the confidence interval of the initial definition. All results remained significant at p=<0.05 and provides evidence that temporal autocorrelation did not impact model robustness. See S6 Figure for the full results of the sensitivity analysis.

### Percentage Attributable Fraction

Based on the randomly resampled IRR values (1,000 samples) from the model results (3.6 for Nigeria and 2.6 for the DRC), the onset of a conflict event during epidemiological week 1 in 1997 to week 20 in 2020, was attributable to 19.7% (95%CI = 18.2%-21.2%) of cholera outbreaks in Nigeria and 12.3% (95%CI = 10.2%-14.4%) for the DRC.

## Discussion

Conflict events increased the rate of cholera outbreaks by 3.6 times in Nigeria and 2.6 times in the DRC. The percentage of cholera outbreaks attributable to the conflicts during the period of 1997 to 2020 (1,220 continuous weeks) was 19.7% for Nigeria and 12.3% for the DRC. The states/provinces with the highest increased risk were Kebbi, Nigeria at 6.9 times and Kasaï-Oriental, the DRC at 7.3 times. This showed that in some states/provinces, the effect of conflict was much greater than the national level.

The sensitivity analysis evaluating the effect of lag showed a decrease in effect as the weeks progressed, with some states/provinces seeing a plateau or increase around 6 weeks after the event. The decrease with the lag duration may be a” diluting” effect, as the probability of an outbreak will increase across a longer period. The states/provinces that increased after week 6, were often those with the strongest initial effect, especially in the DRC. The larger initial effect having a longer lasting impact may potentially be due to conflict severity. The IRR values remained above 1 (2.08 Nigeria and 1.5 for the DRC) at 10 weeks after the conflict, providing further evidence of a long lasting impact of conflict.

States/provinces with the greatest increased rates of cholera often coincided with areas of high conflict. This provides further evidence to the hypothesis that conflict may be a driver of cholera in Nigeria and the DRC. States/provinces surrounding high conflict areas were also highly significant areas (e.g., Abia, Ogun, Osun, Maniema, and Tanganyika), showing a potential spill-over effect. The states/provinces here were studied independently but a possible explanation may be people fleeing areas of conflict or a cholera outbreak to neighbouring states, as displacement is a known risk factor for disease outbreaks (*9*). This is especially important for cholera, as a large proportion of people will be asymptomatic but can still shed the pathogen into local reservoirs, which people may use as drinking water due to a lack of alternatives (*33*).

Cholera outbreaks can be explosive and self-limiting, due to the high number of asymptomatic individuals, diluting the susceptible pool (*33*). This potentially explains why the impacts of conflict on cholera was seen just one week after the event. The incubation period of cholera is short (*34*), making the effect within the first week found here biologically possible for the pathogen and a realistic timeframe for elevated exposure to manifest in cases. Other examples of cholera cases emerging within the first week after an adverse event include Cyclone Thane hitting the Bay of Bengal (*35*), water supply interruption in the DRC (*36*) and Cyclone Aila in West Bengal (*37*). This provides further evidence of the need for quick and effective aid in humanitarian crisis to avoid outbreaks and reduce mortality (*38*).

Healthcare facilities can suffer in periods of conflict and cholera outbreaks can overwhelm systems, a potential cause of the relationship between conflict and cholera shown here. Care can be inaccessible because of direct infrastructure damage or difficulties getting to the facilities due to impromptu roadblocks (*39*). Supplies may be stolen and/or unable to be delivered, including oral rehydration solution (ORS), pathogen-sensitive antibiotics and oral cholera vaccines, which are important for cholera outbreak control and mortality (*40*). Finally, safety is a serious issue, both for healthcare workers and patients and non-governmental (NGO) organisations can withdraw from these areas, citing an inability to ensure the safety of their staff (*41*). Steps need to be taken globally to reduce violence against healthcare, such as using active clinical management for all patients to enhance the acceptance of pathogen-specific treatment centres (*42*).

Conflict has the potential to worsen pre-existing vulnerabilities, which can exacerbate poverty, another potential cause of the effect of conflict on cholera. The impacts of poverty can be far reaching and is a known risk factor for cholera (*4,43*), along with other diseases (*44*). For example, poor urban settlements have faced the brunt of outbreaks including Zika, Ebola, typhoid, and cholera, due to crowding and poor access to WASH (*45*). Conflict can result in loss of possessions, habitual residence, and an inability to find employment, reducing income generation, savings and financial backstops (*13*). In times of worsening poverty, people may not be able to afford healthcare and basic medical supplies, especially in vulnerable groups. This disruption to daily life can cause many more deaths than direct battlefield fatalities and leads to stagnation in development (*46*).

Although WASH and poverty were not directly evaluated, a lack of WASH facilities is likely to have contributed to the positive relationship between cholera and conflict found here. Conflict events can lead to disruption in sanitation and hygiene and adverse events can act as catalysts in the interaction of contaminated water and the human population (*3*). Displacement from conflict can cause issues in accessing WASH (e.g., latrine access, soap availability) and several displacement camps have seen rapid cholera outbreaks, including the DRC after the Rwandan genocide in 1994 (*2*). If people are displaced due to conflict, this may result in the use of water contaminated with toxigenic strains of *Vibrio cholerae* because alternatives are lacking, leading to outbreaks.

A potential limitation is the plausible existence of multiple causal pathways, leading to misclassification due to time-variant confounders. Examples include a conflict event in an adjacent geographic area being causally linked to the conflict event in the current geographic area or the presence of waterbodies, which are considered fundamental in cholera transmission (*47,48*). Understanding additional environmental factors including seasonal weather changes and the pre-existing vulnerabilities discussed are very important, although beyond the scope of the methods used here which investigate conflict in isolation.

Underreporting, overreporting and a reporting lag may have impacted the degree of effect found here. Underreporting is a significant issue in global cholera and conflict estimates, due to asymptomatic cases, disincentives to report and logistical issues (*29,49*). Cholera surveillance is difficult in conflicts, due to displaced populations and security issues. Additionally, our methodology may have resulted in a classification bias, underestimating the effect of conflict on cholera. If a cholera outbreak was “imported” from a neighbouring state/province (spatial autocorrelation), this would be classified as a genuine, autochthonous event, which would likely be non-differential (likely to happen during an exposed or non-exposed period). Alternatively, during times of conflict health surveillance can be enhanced by the government and/or NGOs. Reporting delay is another potential problem and some national reporting delays, have been found to range from 12 days for meningococcal disease to 40 days for pertussis (*28*).

The SCCS model is a case-only approach and analysing cases only, instead of the corresponding complete cohort, translates into a loss of efficiency. However, previous work has shown that this loss is small, especially when the fraction of the sample experiencing the exposure is high (which is the case here). Moreover, this loss of efficiency must be weighed against a better control of time-invariant confounders. Previous examples illustrated that the SCCS design is likely to produce more trustful results than the corresponding cohort analysis, especially when a strong residual confounding bias is likely (*30,31*).

The severity or intensity of both the conflict and cholera outbreaks were not evaluated here, as a binary variable was used. Conflict severity is complex, far-reaching and challenging to measure. Making assessments and assumptions of how a conflict event impacts a health outcome is difficult and may involve oversimplifications. Although beyond the scope of this work, conflict severity is an important area of future qualitative research with those working in a variety of different organizations in the conflict-affected areas.

Despite the limitations of conflict and cholera data, the data used here are to the highest standard currently available and has been used by several other studies, making the research comparable (*11,12*). Additionally, several methods of validating the cholera data were used (*26*). Creating partnerships with those working on the ground and exploring more sensitive data options is an area of future research. Additional methods to account for data limitations included setting both the event and the exposure to a binary outcome to reduce the impacts of severity and using a weekly instead of daily temporal scale to account for delays.

In summary, our analysis shows a clear relationship between cholera and conflict in both Nigeria and the DRC, with conflict increasing the rate of cholera by up to 7.3 times in some states/provinces. The flexibility of SCCS and conditional logistic regression models make future work evaluating different diseases, countries and additional risk factors relatively simple. Cholera risks are likely multi-factorial and complex but sufficient and rapid support, along with enhanced efforts to build community trust can reduce this access risk. Finding conflict resolution and addressing pre-existing vulnerabilities (poverty, healthcare and WASH) should be the main priority. By reducing these vulnerabilities, communities will have greater resources to adapt and reduce vulnerabilities both in times of conflict and peace.

## Data Availability

All data used here are complied from publicly available data sources and the full data is available from: https://github.com/GinaCharnley/cholera_data_drc_nga Conflict data is available from the Humanitarian Data Exchange: https://data.humdata.org

## Acknowledgements

This work was supported by the Natural Environmental Research Council [NE/S007415/1], as part of the Grantham Institute for Climate Change and the Environment’s (Imperial College London) Science and Solutions for a Changing Planet Doctoral Training Partnership. We also acknowledge joint Centre funding from the UK Medical Research Council and Department for International Development [MR/R0156600/1]. We thank the organisations who published and those who collected the data that was used here, including the Nigerian Centre for Disease Control and Minstère de la Santé RDC. Finally, we acknowledge and thank Heather Whitaker (Open University) and Yonas Weldeselassie (University of Warwick) for their assistance and advice in the SCCS methodology.

## Author Bio

Gina Charnley is a Postgraduate Researcher in the School of Public Health at Imperial College London. She is currently on the Science and Solutions for a Changing Planet DTP through the Grantham Institute and her PhD focuses on infectious disease outbreaks during and after disasters such as natural hazards and conflict and how climate change may alter these outbreaks.

## Supporting information

### S1 Information. Cholera case definitions according to the Nigerian Centre for Disease Control and the Ministère de la Santé Publique de la République démocratique du Congo

#### NCDC

Suspected case: Severe dehydration or death from acute watery diarrhoea in a patient aged 5 years or more. In an epidemic situation: A suspected case in any person aged 5 years or more with acute watery diarrhoea with or without vomiting.

Confirmed case: A suspected case in which *Vibrio cholerae* O1 or O139 has been isolated in the stool.

#### RDC Ministère de la Santé

Suspected case: Severe dehydration or death following acute watery diarrhoea in a patient aged 5 years or more. In an epidemic situation: Acute watery diarrhoea with or without vomiting in a patient aged 1 year or more.

### S2 Information. Dataset information

Cholera data was compiled from a range of publicly available sources (WHO’s disease outbreak news, ProMED, ReliefWeb, WHO’s regional office for Africa weekly outbreak and emergencies, UNICEF cholera platform, EM-DAT, the Nigerian CDC and a literature search) in both English and French. A data charting form was used to enable a dynamic data entry process and collected data on date, geographic location, cases, deaths, hospitalisations, fatality rates, gender, age, oral cholera vaccinations, risk factors, aid and the source of the report. Data spanned from 1971-2021 for Nigeria and 1978-2021 for the DRC on a daily temporal scale and was provided at the finest spatial scale possible.

Conflict data was provided by the United Nations Office for the Coordination of Humanitarian Affairs’s Humanitarian Data Exchange (HDX, 2020). The data included sub-national conflict events for both countries on a fine spatial scale, given to the exact location in longitude/latitude. This was reported on a daily temporal scale and spanned from 1997 to 2020. The data was also categorised by event type which included battles, explosions, protests, riots, strategic developments and violence against civilians. This was further sub-categorised within these groups and reported number of fatalities.

The study period was selected as Jan 1997 to May 2020, as these were the first and last reports in the conflict data. The spatial granularity of the analysis was to administrative level 1 (states for Nigeria and provinces for the DRC) and all data points that were reported on a finer spatial scale were attributed to the upper level. To be included in the analysis, the state/province had to report both outbreaks and conflicts during the study period, therefore 22 provinces were included for the DRC and 36 states for Nigeria.

### S3 Information. Sensitivity analysis

Alternative exposure end points to identify the effect of lag. Five alternative exposure periods were tested from the original exposure period (1 week after the onset of exposure, lag 1) and were named lag periods due to the potential lag effect from conflict onset to cholera outbreaks, these included:

1. Lag 2 - Week of conflict onset + 2 weeks
2. Lag 4 - Week of conflict onset + 4 weeks
3. Lag 6 - Week of conflict onset + 6 weeks
4. Lag 8 - Week of conflict onset + 8 weeks
5. Lag 10 - Week of conflict onset + 10 weeks

The sensitivity analysis was run on both a national and sub-national level and S1 and S2 Figs show additional swimmer plots of lag 10 and line plots of the temporal trends.

### S4 Information. Equations used to calculate the Percentage Attributable Fraction

First the number of outbreaks attributable to conflicts, *A*_*i*_, for each province *i*. Is estimated using the formula:

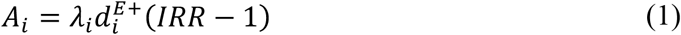

Where 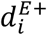 is the total duration of conflict exposure for the province *i* (if no conflict in province *i*, thus 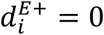), *λ*_*i*_ is the rate of outbreak occurrence in a Poisson process in the absence of conflict, and IRR is the incidence rate ratio associated with exposure to conflict. With 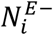 being the number of outbreaks observed in the province *i* during the un-exposed period and *T* being the total period of observation, an estimator of *λ*_*i*_ is 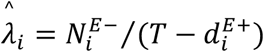, which leads to:

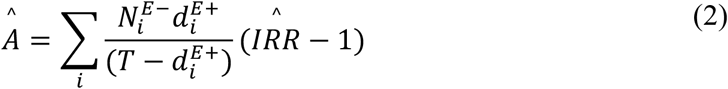

Based on 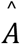 and *N*, the total number of outbreaks observed, we can easily obtain the equivalent of the population attributable fraction, *PAF*, which corresponds to the proportion of the total number of outbreaks in both countries that are attributable to conflicts (this is equivalent to the PAF obtained in classical epidemiological studies, but here population refers to the “population of provinces”):

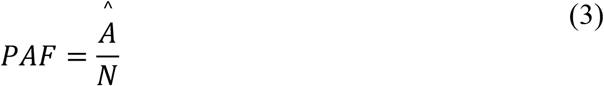

### S5 Information. Excluded events

States/provinces removed as they did not report conflict and cholera in the study period (1997-2020).

Democratic Republic of Congo:

- Haut-Uele - 629 conflict events removed
- Kasaï-Central - 234 conflict events removed
- Lomani - 101 conflict events removed
- Tshuapa - 70 conflict events removed

Nigeria:

- Imo - 239 conflict events removed

**S1 Table.**
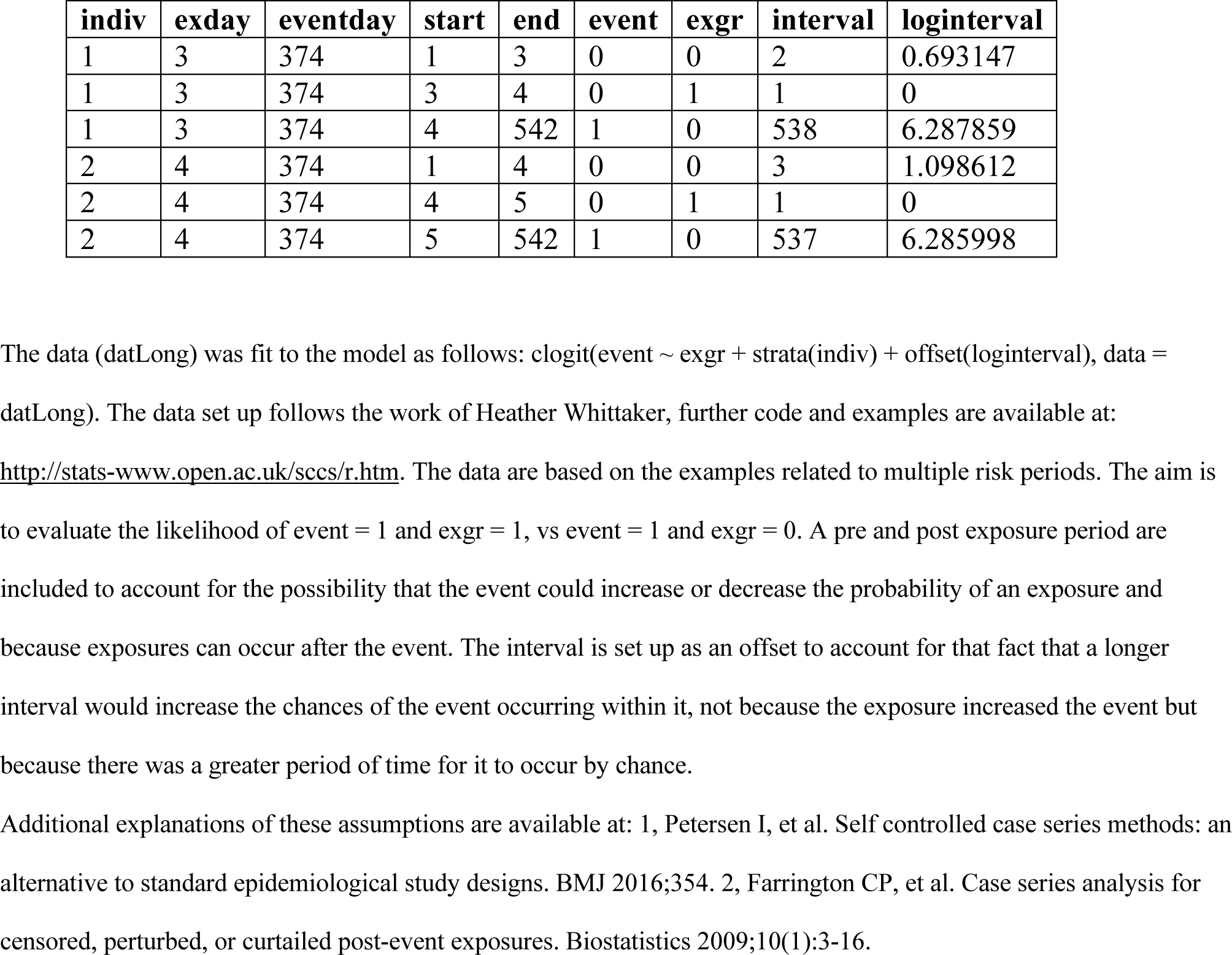
The layout of the pseudo-dataset dataframe fitted to the model. Each event and exposure are given a reference number (indiv).

**S1 Figure.**
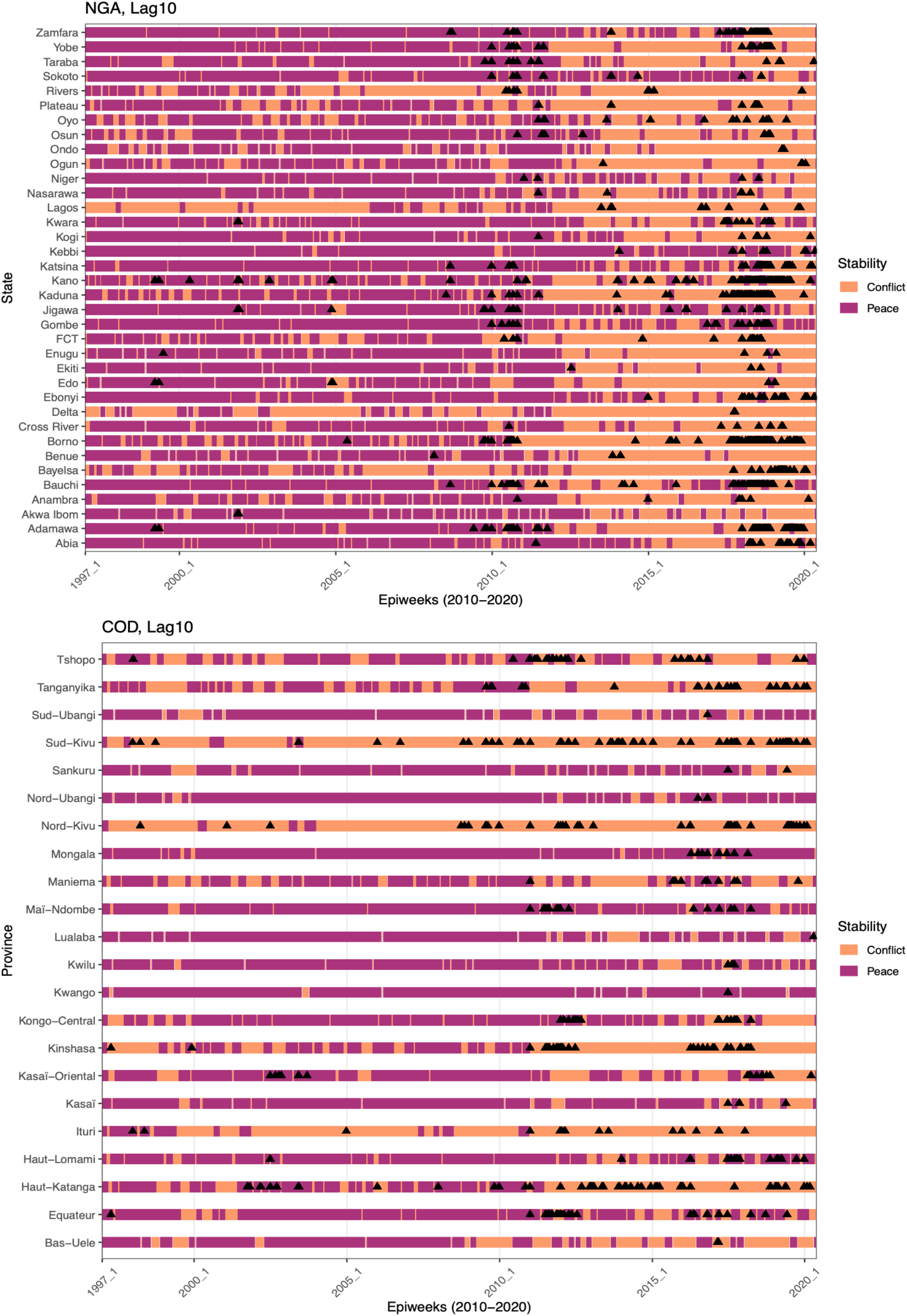
Swimmer plots showing the conflict dataset for lag 10 in the sensitivity analysis. In relation to outbreaks (black triangles) for Nigeria (NGA) and the Democratic Republic of Congo (COD).

**S2 Figure.**
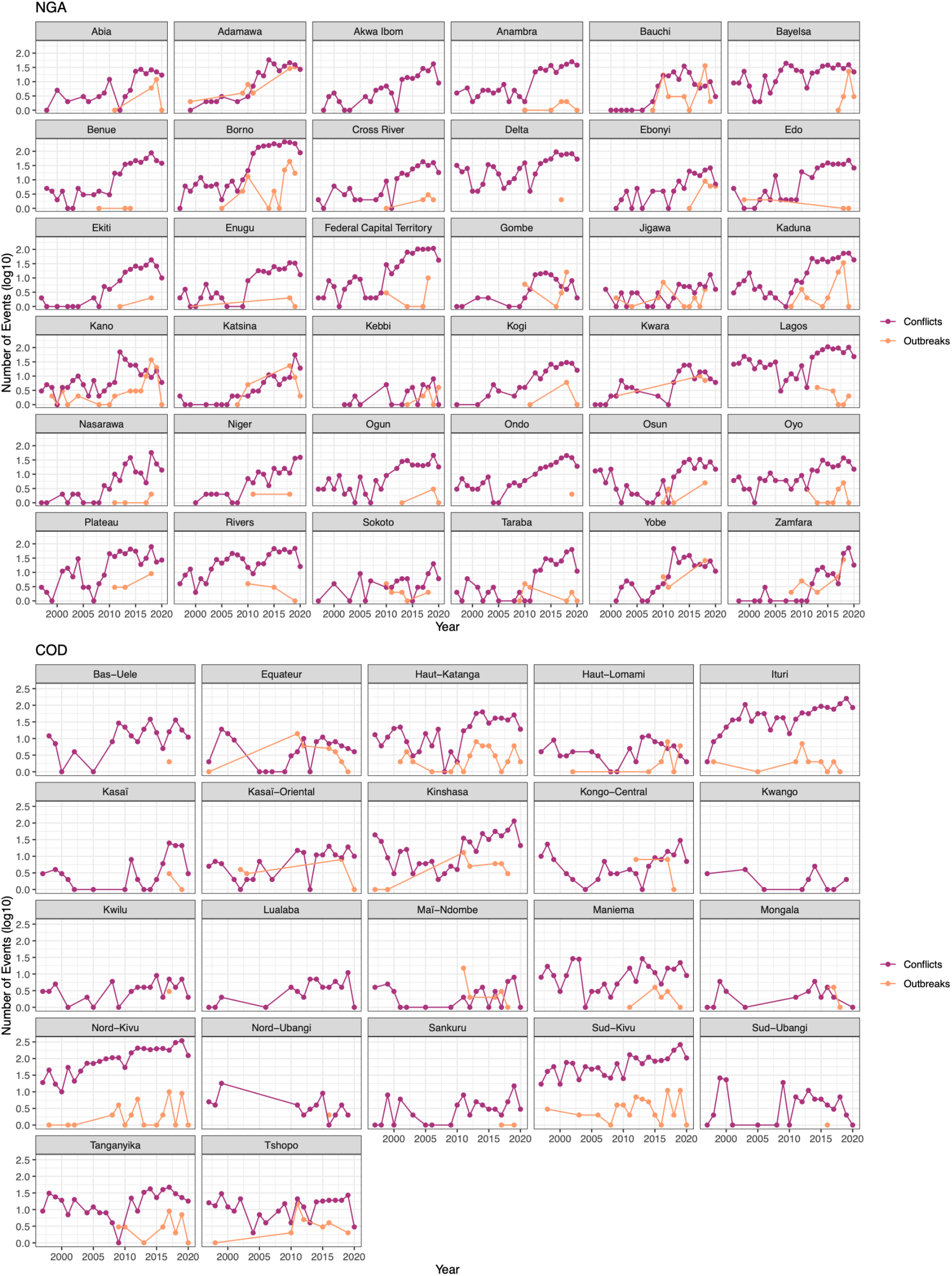
Number of outbreak (orange) and conflict (purple) events by year in Nigeria and the Democratic Republic of Congo over the full study period.

**S3 Figure.**
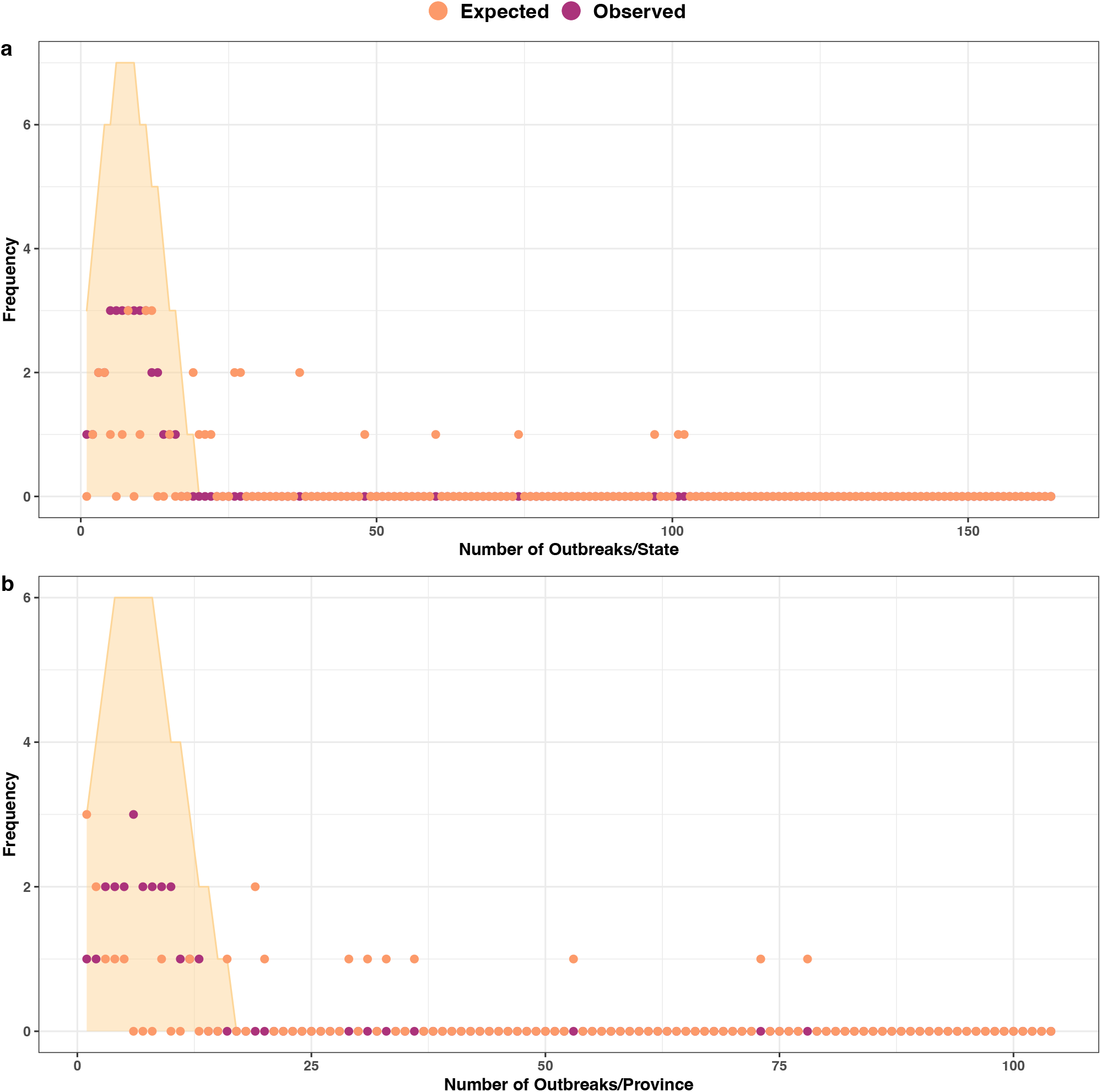
Poisson probability distribution fit to the outbreak data. The simulated counts were obtained from 10,000 random realizations of a Poisson process of rate λ = number of total national outbreaks/number of states or provinces, for **a**, Nigeria and **b**, the Democratic Republic of Congo. Expected values are the median simulated counts from the distribution with a 95% confidence interval.

**S4 Figure.**
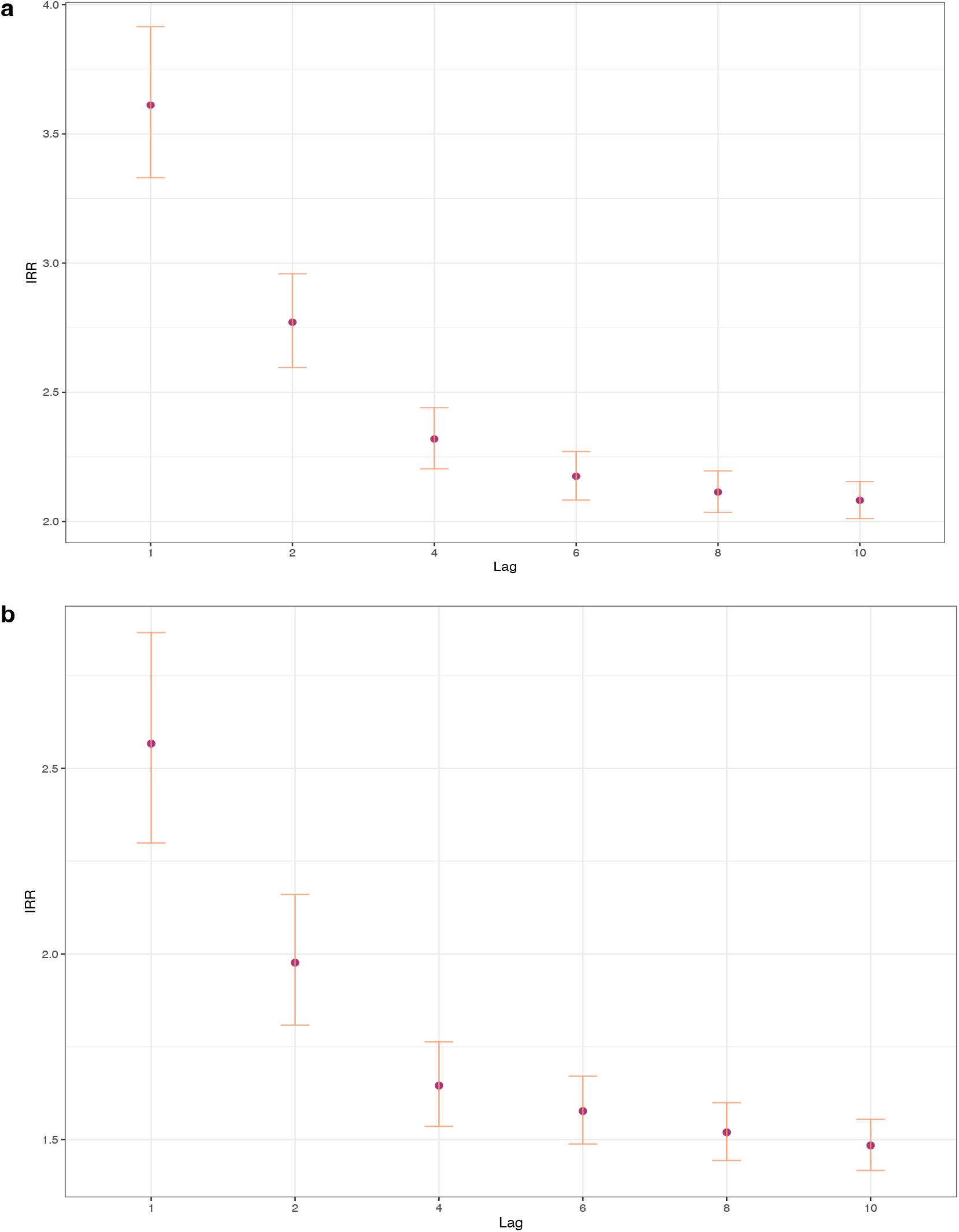
Results of national lag period sensitivity analysis. Incidence rate ratio (IRR) for the effect of exposure to conflict within 1, 2, 4, 6, 8 and 10 weeks of the event and cholera for **a**, Nigeria and **b**, the Democratic Republic of Congo. Only results that were significant at the threshold p=<0.05 are plotted here. From week 1 to week 10 the risk decreased from 3.6 to 2.08 for Nigeria and from 2.6 to 1.5 for the DRC. This suggests that the risk of conflict on cholera is highest soon after the event but remains a detectable association albeit at a lower level for potentially a long period of time after the event.

**S5 Figure.**
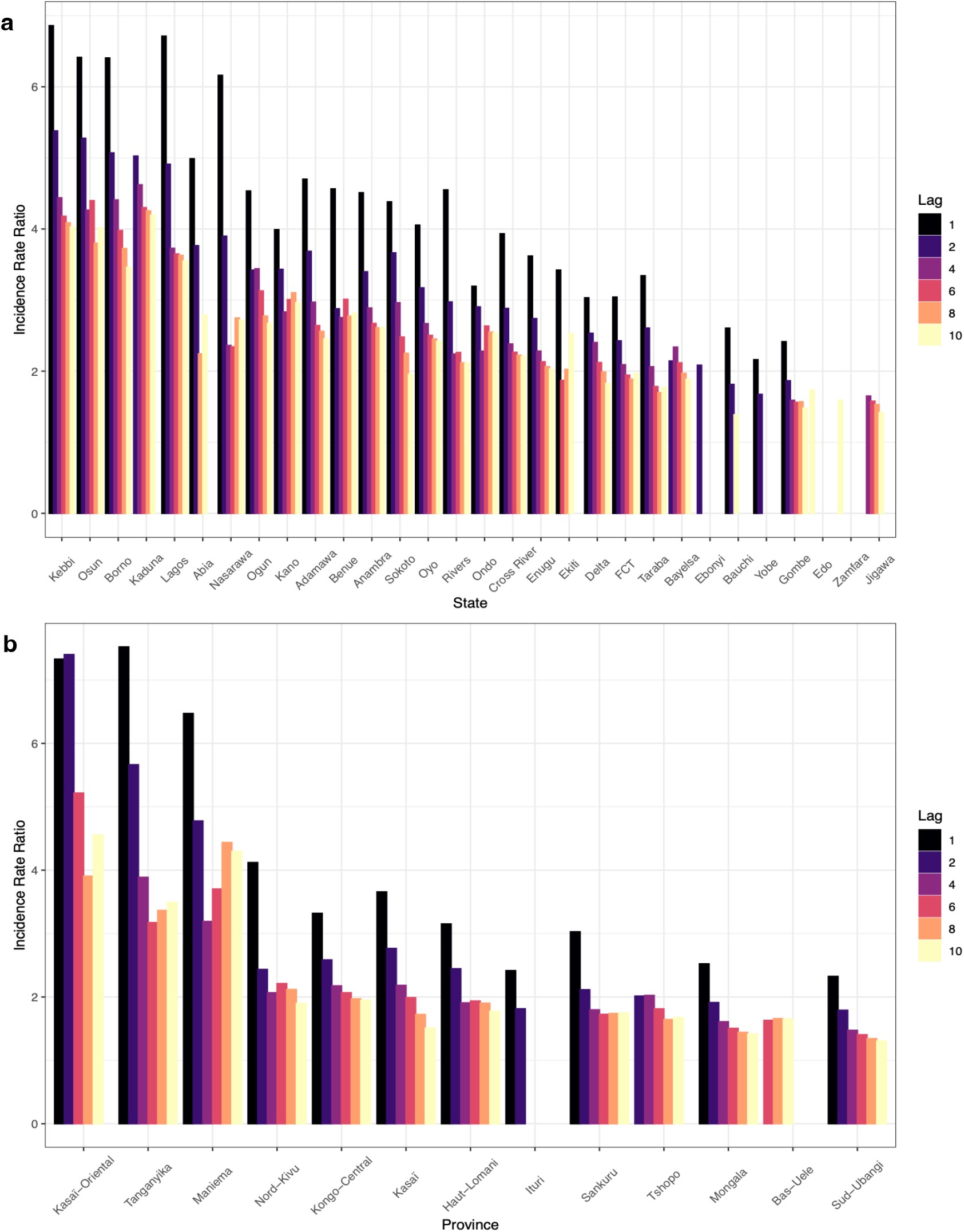
Results of subnational lag period sensitivity analysis. Incidence rate ratio (IRR) for the effect of exposure to conflict within 1, 2, 4, 6, 8 and 10 weeks of the event and cholera at administrative level 1. For **a**, Nigeria and **b**, the Democratic Republic of Congo. Only results that were significant at the threshold p=<0.05 are plotted here. Thirty Nigerian states and 13 DRC provinces were found to be significant for at least one of the lag periods and the most significant states predominately followed the trends of the national analysis. Values ranged from Kebbi at 6.9 to 4.0 times increased risk of cholera, to Gombe at 2.4 to 1.5.

**S6 Figure.**
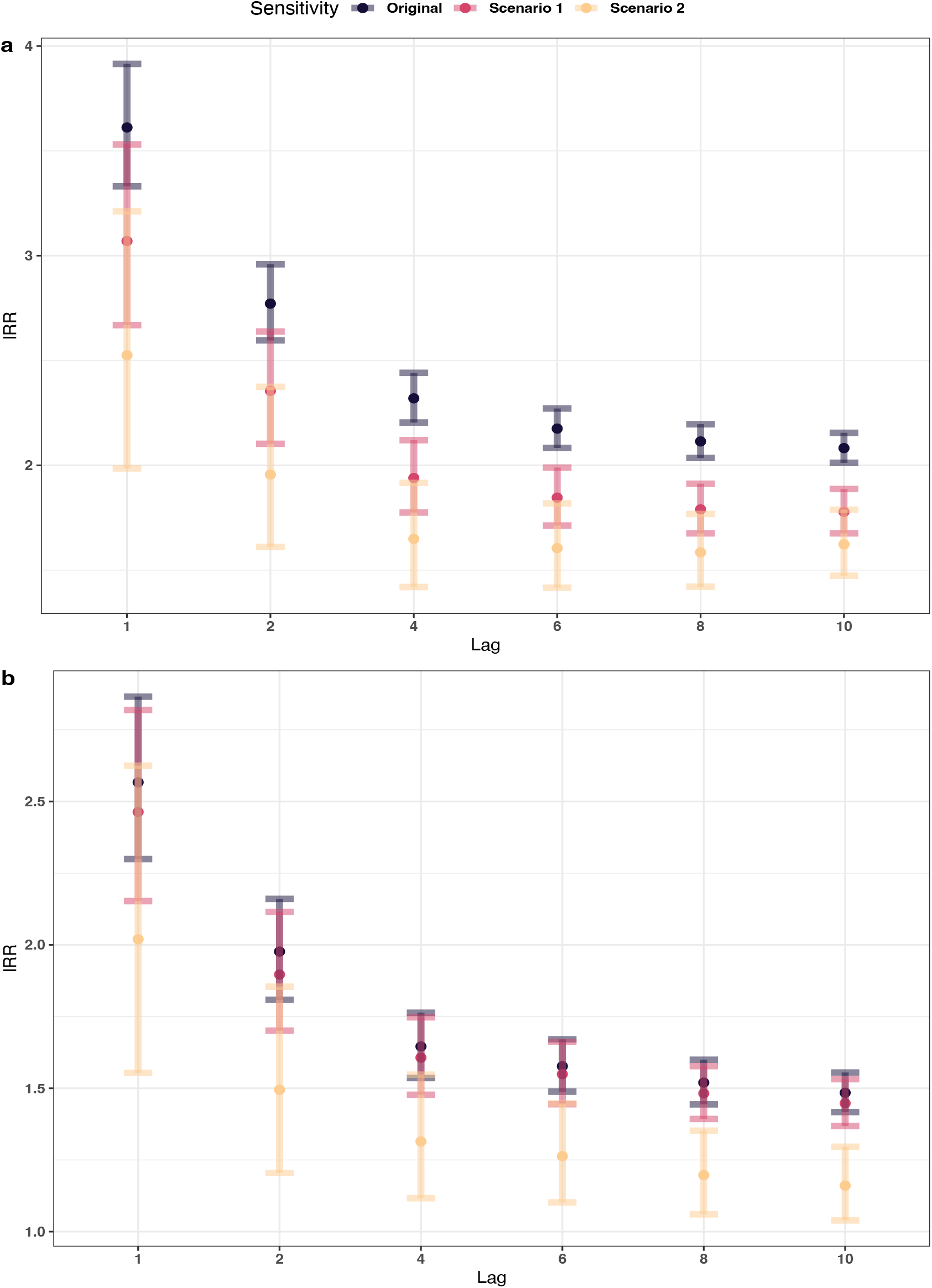
Results of outbreak definition sensitivity analysis. Incidence Rate Ratio (IRR) values and 95% confidence interval for **a**, Nigeria and **b**, the Democratic Republic of Congo for Scenario 1 removing all outbreaks within 2 weeks of each other (10 days shedding + 5 days incubation) and Scenario 2 removing all outbreaks within 6 months of each other. Both alternative scenarios are compared against the “Original” analysis, using the outbreak definition of 1 or more cholera cases being reported in a specific week.

